# Anaesthesia provision challenges in public hospitals of Pakistan’s Punjab province: a qualitative study of expert perspectives

**DOI:** 10.1101/2023.04.13.23288520

**Authors:** Sumbal Shahbaz, Rubeena Zakar, Natasha Howard

## Abstract

**Background:** Anaesthesia delivery in Pakistan remains limited to conventional intraoperative procedures, with research showing ongoing challenges in quality and resourcing. We aimed to identify systemic challenges in the delivery of quality anaesthesia services for surgical support in Pakistan’s Punjab province.

**Methods:** This qualitative study included 22 semi-structured interviews with purposively selected anaesthesia system experts in Punjab province, including heads of teaching hospital anaesthesia departments, healthcare commission representatives, and health department officials. We analysed data thematically, using deductive and inductive coding.

**Results:** We identified three themes of anaesthetist recruitment and retention, quality-of-care and in-service training, and discrepancies between specialities, describing major challenges experienced within the speciality. Findings indicated that workforce shortages and maldistribution, insufficient in-service training and standards, inadequate equipment maintenance, and lack of anaesthesia representation in decision-making compromised anaesthesia provision quality and safety.

**Conclusions:** Improving anaesthesia provision in Punjab would require increasing physician and non-physician anaesthetist numbers and rotation to peripheral postings, strengthening training quality, and ensuring availability of minimum essential equipment and supplies. To achieve essential anaesthesia provision standards, policy interventions are needed to, for example, balance anaesthesiologist and surgeon numbers, require that anaesthesiology graduates work a few years in-country (e.g. scholarship bonds), ensure in-service training attendance for skills updates, and implement quality assurance standards for equipment and supplies.

**HIGHLIGHTS:** *What is already known on this topic?:* Existing research on anaesthesia in lower-income economies focuses on provision discrepancies and capacity measurement of HIC-partnered interventions. However, managerial and frontline challenges that weaken quality anaesthesia provision in countries such as Pakistan, and thus affect global surgery indicators, are largely unexamined.

*What this study adds:* This study is the first to highlight ongoing challenges within the anaesthesia delivery system in Punjab province as experienced by senior practitioners and health officials, thus contributing to the knowledge base on anaesthesia provision challenges in lower-income economies.

*How this study might affect research, practice, or policy:* Findings show the urgent need to increase recruitment, retention, and peripheral distribution of physician and non-physician anaesthetists along with developing clear national legislation and practice guidelines for standardised quality of anaesthesia care and raising the public profile of anaesthesia in Pakistan.

## INTRODUCTION

Anaesthesia can generally be categorized as insensitivity to pain, particularly by induction of synthetically prepared gases or inoculation of drugs prior to surgical procedures, and typically comprises of hypnosis, amnesia, and analgesia along with muscle paralysis (1). Anaesthesia provision has evolved beyond the operating theatre in the past twenty years, including for management of critical pain and palliative care (2). Modern anaesthetists are skilled in cardiopulmonary resuscitation, basic and advanced life support, and catastrophe management, with vital roles in trauma centres, critical care units, pain clinics, and resuscitation units globally (3).

While data support the cost-effectiveness and improved outcomes of medical procedures conducted with trained anaesthesia personnel, the value of anaesthesiology is not widely recognised in lower-income countries (10). Lower-income countries such as Pakistan deal with multiple misconceptions related to anaesthesia cost and outcomes that affect patient willingness to obtain surgical and anaesthetic interventions and therefore negatively affect patient outcomes and population health (8). Despite significant advancements in anaesthetic techniques and quality coupled with improved surgical success rates, there is insufficient public awareness of anaesthesia’s importance in healthcare delivery and it is still considered a hidden specialty (9).

In Pakistan, surgeries are only provided in secondary and tertiary health facilities, so anaesthesia professionals only work in Tehsil (township), District, and teaching hospitals (4) Anaesthesiologist are recruited in two cadres: (i) Academic consultant, with Master of Science (MS) or Fellowship training for the College of Physicians and Surgeons (FCPS) in anaesthesia and clinical, teaching, research and administrative duties; and (ii) Clinical, with Diploma (DA) or Membership of the College of Physicians and Surgeons (MCPS) in Anaesthesiology, only providing clinical services (5). Teaching hospitals train residents for degrees in anaesthesia including the two-year DA or MS in Anaesthesiology, or two-year MCPS or 4/5-year FCPS. A few institutes also offer non-physician anaesthesia training. While anaesthesia procedures are well defined by the Healthcare Commission (HCC), anaesthesia delivery is generally restricted to conventional surgical procedures (2). Thus, anaesthesia is limited to a few hospitals and subspecialties and has not improved sufficiently, with disparities increasing as the population grows (6, 7).

Bashford & Vercueil noted that most literature on anaesthesia provision in low and middle-income countries (LMICs) is published by high-income country (HIC) authors (11) and is predominantly focused on capacity measurement, evaluation of HIC-partnered interventions (12), or quality-improvement initiatives supported by international donations or equipment provision (13, 14). Locally conducted anaesthesia research is thus uncommon in LMICs, due to lack of research capacity, funding, and interest, worsened by continued emigration and ‘brain drain’ of both physician and non-physician anaesthetists from LMICs to HICs (15-17).

This study thus aimed to explore systemic challenges in the delivery of quality anaesthesia services in public-sector health facilities in Punjab province, Pakistan. Objectives were to: (i) identify perceived challenges; (ii) synthesise major systematic issues; and (iii) consider potential anaesthesia delivery system improvements of relevance in Punjab and potentially elsewhere.

## METHODS

### Study design and research question

We chose a qualitative approach, as described by Teherani et al, to gain insight into organisational and systemic issues in anaesthesia delivery through examining perspectives of expert key informants in Punjab province (18). Our research question was: “What do anaesthesia experts identify as the key challenges in provision of quality anaesthesia services in public-sector hospitals in Punjab province?”

### Sampling and recruitment

We purposively sampled experienced anaesthesia services stakeholders within the Punjab health system. We first identified three stakeholder domains able to provide insight on strategic challenges for this speciality: (i) heads of anaesthesia departments in teaching hospitals; (ii) HCC representatives; and (iii) health department officials. Eligibility criteria were having served five or more years within their respective domains, having contributed to anaesthesia policymaking and implementation, and being over age 18 and able to be interviewed in Urdu or English. SS contacted 22 potential participants through their official phone number or email, of whom none refused to participate. We continued recruitment until we considered we had achieved data saturation, i.e. repetition of relevant information (19).

### Data collection

#### Tool development

We developed a semi-structured interview guide based on literature review and expert opinion (20) comprising six topics, namely demographics (e.g. age, position, years of experience), routine anaesthetic care, anaesthesia workforce distribution and maldistribution, recruitment and retention problems, government actions to address issues, interviewee actions to address issues. The guide allowed for discussion of deductive concerns, probing of new or interesting elements, and emergence of unexpected sub-topics.

#### Interviewing

SS contacted potential interviewees on their work phone to explain the study and invite participation. SS conducted in-person semi-structured interviews in Urdu during June-September 2021, after audio-recording verbal informed consent, as government employees are generally unwilling to sign consent forms in case they might pose a job risk. Interviews took approximately minutes and were audio-recorded with participant consent. One did not agree to audio recording, so SS took detailed notes. SS translated and transcribed notes and audio files into English, which were then checked by RZ. SS and RZ determined data saturation was achieved when no new concepts emerged from interviews. We ensured participant anonymity and confidentiality by allowing participants to choose interview times and locations, using identification codes instead of names on all outputs, destroying audio files after transcription, and storing transcripts in a password-protected hard drive only accessible by researchers.

### Analysis

SS analysed transcript data thematically in English, as described by Braun & Clarke (21). This consisted of data familiarisation, generating initial codes, searching for themes, reviewing, defining, and naming themes with NH, and writing up the analytic narrative and extracted quotes.

### Patient and public involvement

Patients and public were not involved in research design or implementation, as our focus was on experiences and perspectives of anaesthesia providers in Punjab.

### Ethics

The University of the Punjab Department of Public Health Doctoral Program Committee provided study approval (D/154/ISCS; March 18, 2021).

## FINDINGS

### Participant characteristics and themes

Table 1 shows characteristics of the 22 interviewees included. Ten (45%) were anaesthesia department heads, 7 (32%) were health department officials from specialist, primary, and secondary care, and 5 (23%) were from the Punjab healthcare commission. Most were men (18; 82%) and aged 45-55 (11; 50%). All were significant anaesthesia stakeholders in Punjab and active in health legislation for over 5 years.

We included three overarching themes: (i) recruitment and retention; (ii) quality-of-care challenges; and (iii) discrepancies between specialities, and included inductive sub-themes within each theme as appropriate.

**Table 1.** Participant characteristics.

| ID | Role | Gender | Age range (years) | Experience |
| --- | --- | --- | --- | --- |
| HA1 | Anaesthesia Department Head | Male | 45-55 | 15 years |
| HA2 | Anaesthesia Department Head | Male | 45-55 | 15 years |
| HA3 | Anaesthesia Department Head | Male | 55-65 | 25 years |
| HA4 | Anaesthesia Department Head | Female | 55-65 | 15 years |
| HA5 | Anaesthesia Department Head | Male | 55-65 | 28years |
| HA6 | Anaesthesia Department Head | Male | 55-65 | 30 years |
| HA7 | Anaesthesia Department Head | Female | 55-65 | 30 years |
| HA8 | Anaesthesia Department Head | Female | 55-65 | 24 years |
| HA9 | Anaesthesia Department Head | Male | 45-55 | 18 years |
| HA10 | Anaesthesia Department Head | Male | 45-55 | 19 years |
| HC1 | Healthcare Commission | Male | 55-60 | 24 years |
| HC2 | Healthcare Commission | Male | 45-55 | 9 years |
| HC3 | Healthcare Commission | Male | 45-55 | 6 years |
| HC4 | Healthcare Commission | Male | 45-55 | 8 years |
| HC5 | Healthcare Commission | Male | 35-45 | 9 years |
| HD1 | Health Department | Male | 35-45 | 5 years |
| HD2 | Health Department | Male | 35-45 | 8 years |
| HD3 | Health Department | Female | 35-45 | 6 years |
| HD4 | Health Department | Male | 45-55 | 9 years |
| HD5 | Health Department | Male | 45-55 | 5 years |
| HD6 | Health Department | Male | 45-55 | 7 years |
| HD7 | Health Department | Male | 45-55 | 6 years |

### Recruitment and retention

Participants described seemingly inefficient human resource processes, in which enough anaesthesia students were trained but neither recruited nor retained in the public sector. We identified four inductive sub-themes related to recruitment and retention: (i) shortages of qualified anaesthetists; (ii) geographic maldistribution; (iii) consultant anaesthetist emigration; and (iv) female anaesthetist dropout.

#### Shortages of qualified anaesthetists

Many participants described slow and complex public sector recruitment processes, which contributed to anaesthesia staffing shortages.

> *“[If] I want a resident who passed exams to work with me in my hospital, the procedure is so difficult! I would write to the principal, who in turn would write to the health secretory. He would analyse seats, advertise them, organise a selection committee, and a lot of other protocols, which would take 3-6 months. Meanwhile, the consultant who just passed out would grasp any new opportunity instead of waiting jobless for months for the desired seat*.*” (HA10)*

Almost all participants reported significant shortages of qualified anaesthetists, particularly experienced ones able to train medical officers (MOs) and postgraduates in teaching hospitals. This overburdened available faculty members and senior registrars and compromised quality of both training and patient care.

> *“Consultant anaesthetists in government facilities are scarce. For every 4 to 5 seats we just have one consultant, who must train postgraduates including DA, MS, FCPS Anaesthesia, MCPS Anaesthesia students along with undergraduate programs, has to attend faculty meeting, work on rosters, give lectures, make and conduct exams, vivas, and supervise theatre lists of more than 10 specialities as well*.” (HA3)

Several described the burden in teaching hospitals as aggravated by increased trainee numbers and fewer teaching staff. Potential reasons were unclear, with some suggesting this was either an individual failing in that anaesthetists were less willing to serve their country or a structural failing in that anaesthesia had been overlooked as surgical needs expanded.

> *“With time, the field of surgery has manifestly improved. Every hospital has numerous surgical specialities. However, anaesthesia must accommodate everyone, as no surgery is possible without anaesthesia. So, shortages remain the same due to lower ratios of anaesthetists compared with multiple surgical specialities*.” (HA8)

#### Geographic maldistribution

Most participants identified anaesthesia workforce maldistribution as a major issue, especially in south and west Punjab where anaesthetist numbers were minimal. Reasons suggested were complex, but primarily related to insufficient qualified anaesthetists to fill urban posts, making them unlikely to seek rural or remote posts.

> “*Surgeons, ENT specialists, gynaecologists opt for remote areas because seats in cities are full and they have fewer opportunities to move abroad. Anaesthesia consultants have immediate opportunities abroad, and those who could, join hospitals in bigger cities. That is why anaesthetists are so few in peripheral areas*.*” (H9)*

A second reason several described was the high anaesthesia workload in peripheral facilities, reinforced by insufficient staff.

> *“In tertiary care a consultant gets one call per week, but in DHQ/THQ [District/Tehsil Headquarters Hospital] a consultant is on call 24/7 […]. He must work in theatre and do administrative work due to lack of workforce, with fewer chances of private practice. So, nobody wants to get in a mess by going to the periphery!” (HD2)*

In addition to heavy peripheral workloads, some also described bullying of those not achieving unrealistic performance expectations.

> *“One of my senior consultant friends moved to a THQ hospital, as he was offered a good package and the hospital was near his hometown. However, he had to quit within 2-3 months as he was expected to run two or more tables at once with no medical officer or assistant. On refusing, he was harassed by fellow surgeons and MS [Medical superintendent] who would report he was unwilling to work. So, to maintain his mental health, he quit the job and started private practice in a bigger city. This kind of attitude promotes maldistribution*.*” (HA7)*

Many participants suggested incentives or required rotations were necessary. A health department official suggested peripheral rotation should be a required element of post-graduate anaesthesia practice, to enable anaesthetists to work in multiple settings and help reduce maldistribution and excessive workloads in peripheral areas. However, both requirement and incentive approaches would need careful consideration, as unrealistic expectations were also apparent in incentive structures - as a senior participant explained:

> *“To reduce maldistribution, previous governments tried to give incentives by allowing consultants working in DHQs to go to nearby government facilities in evenings and do cases for 5000 PKR per case or sometimes even more. Their scheme was good, but they didn’t realize anaesthesia is not just intraoperative, you must give preoperative and post-operative care. You just can’t leave a patient after surgery. If a post-operative emergency happens, the blame would be on the anaesthetist if he leaves. Without a proper caregiving system anaesthesia is impossible to administer*.” (HA10)

#### Consultant anaesthetist emigration

Most qualified anaesthetists emigrated due to global anaesthetist shortages and better working conditions outside Pakistan. Qualified anaesthetists did not need additional exams to begin working in higher-income countries and approximately 90% of FCPS consultants reportedly emigrated immediately after graduation every year.

> *“Several of our FCPS trainees pass exams every year and immediately go abroad to work, as they are hired by Saudi, UAE, UK, and Ireland without any additional exam. This could be a good thing personally, but it burdens our system here, as one skilled anaesthetist then has to do the work of 5 people*.*”* (HA2)

Participants described this as higher-income countries freely benefiting from Pakistan’s educational efforts.

> “*We are facing a sheer shortage of anaesthesia workforce for the last 3 decades and it’s all over the world, but this factor is affecting us more as the developed world is drawing our trained staff*.” (HC1)

Reducing consultant emigration was identified as an urgent priority to improve the numbers of skilled anaesthesia professionals in Pakistan. Many suggested bonding anaesthesia students to remain in government service for a few years before being free to leave.

> “Senior *faculty give time and energy to residents. Government also pays them a handsome stipend. It is unjust to complete specialisation and fly off to another country immediately. They must be bound here to work for at least 3 years and train a few more people. Moreover, failing to do so must come with a heavy fine, and penalties such as returning all the stipend they received here or cancellation of basic medical degree*.” (HC2)

Many participants noted this skill exodus was primarily relevant to anaesthesia, hindering any likelihood of reforms. Health department officials reinforced this assertion by suggesting bond legislation would be hard for a poor country such as Pakistan, whose biggest export is its qualified personal, to implement. As one noted, despite complaints no action had yet been taken.

> *“Consultants could be bound by policy introduced by PMC [Pakistan Medical Commission] and implemented by the health department. Unfortunately, the senior doctors just complain but do not actually do what’s required*.*” (HD2)*

#### Female anaesthetist dropout

Most participants indicated that emigration was gendered, and women anaesthetists could significantly benefit the anaesthesia delivery system, if appropriately supported, as they were less likely to emigrate due to familial ties and responsibilities.

> “*Female anaesthesia workforce could be a game changer for our country, as they don’t move abroad (even with excellent packages) due to families here. They could bring a massive improvement in the present condition of the anaesthesia system if provided proper facilities and opportunities here in government and private sector*.” (HD4)

However, many also noted that familial responsibilities were also gendered in Pakistan with women not readily able to work nights and requiring periods of leave for maternity and childcare duties.

> *“Female health professionals are an indispensable part of our system. They work dedicatedly throughout training. However, when 70% of your trainees are female, it becomes difficult to manage with maternity leaves and their hesitation for evening and night duties. We cannot force male doctors to work at night or evening all the time*.” (HA4)

Several participants complained that most anaesthetist trainee seats were occupied by female residents, many of whom stopped working once they married or had children.

> *“Of 100 female consultant anaesthetists, only 20-30% do regular practice, which creates a big gap in quality of anaesthesia provision in both government and private sector*.*” (HD6)*

### Quality-of-care challenges

We identified four related sub-themes affecting anaesthesia quality-of-care: (i) unnecessary referrals; (ii) intraoperative focus; (iii) lack of quality assurance; and (iv) pre-operative and intra-operative training gaps.

#### Unnecessary referrals

Most participants described unnecessary referrals as a major health system flaw, which burdened tertiary healthcare and increased morbidity and mortality. However, suggested reasons differed significantly. Some described it a result of maldistribution and overwork, and others as an excuse not to work.

> *“The same anaesthetists who would refer a case to tertiary care would love to do it in private facilities with even fewer resources to make extra money. Unnecessary referrals are just lack of devotion to the profession and haste to make money*.*” (HC3)*

Anaesthesia was one of the main reasons for referral, as it was not available in most peripheral hospital ICUs. However, some suggested this should not be a barrier.

> “*Not every case requires post-operative ICU, but Districts and Tehsil level hospitals refuse to perform simple laparotomies and hysterectomies due to unavailability of ICU. This is just unwillingness to work, which is not an excuse to play with the patient’s life. Strict actions must be taken to overcome this*.*” (HD4)*

However, HCC participants suggested that infrastructure was a problem and doctors were right to refer.

> *“As per law, in every 10-bed hospital there must be one ICU bed, which we do not have. And in some places, we have ICU but no staff to run it. So sometimes we cannot challenge the concerns of anaesthetists as its infrastructure issues that must be overcome*.*” (HC4)*

Meanwhile, health department participants complained that they could not act on potential problems without sufficient information.

> *“There are rules and doctors could complain about unnecessary referrals, but they don’t in the name of brotherhood and just blame the government. Government can’t work on a problem unless it’s reported*.*” (HD3)*

Other participants suggested interventions, such as mobile ICUs to provide immediate post-operative ventilation and reduce operative referral to tertiary care.

> “*Not every case requires ICU. So, instead of making ICU in every hospital government should make mobile ICU in ambulances, which should be available on request. This way patients could be given immediate post-operative ventilators instead of referring cases to tertiary care…” (HD5)*

All participants recommended clear referral rules, administered by both referring and receiving hospitals, so cases could not be referred for no obvious reasons.

#### Intraoperative focus

Many participants insisted that most work in any hospital required anaesthesia support - pre, intra, or post-operatively, but government hospitals focused on expanding intraoperative anaesthesia care, so anaesthetists seldom provided additional pain management or post-operative care.

> “*In most government facilities, anaesthetists themselves do not know about their responsibilities. Government makes bigger and bigger surgical wards, but no post-op wards […] as they consider anaesthesia just an intra-operative field. So, most government anaesthetists think ‘why should we increase our responsibilities, by going into pain management or post-op care with no extra benefits?’ Instead, they try to give time to private practice…”* (HA4)

Several mentioned that insufficient experienced anaesthetists meant insufficient time to train juniors on expanded anaesthesia care despite the benefits.

> “*This hospital is the only one in Punjab running a pain clinic. No other government hospital does because it requires extra effort, more duty hours, more stress but in turn it not only benefits our terminally ill patients but also improves skills of our residents who could practice it in future*.” (HA6)

Others suggested lack of interest was primarily financial, as neither government nor private sectors offered incentives for expanding preoperative and postoperative anaesthesia.

> “*Government doesn’t offer any bonus on expanding the boundaries, nor does private sector give space to anaesthetists practicing post and preoperatively. A surgeon thinks he can manage post-operative pain, orthopaedic surgeons don’t let anyone deal with joint pains, oncologists think only he can deal with cancer patients*… *When nobody would refer patients to us, why would we take extra pain [to practice additional skills]?”* (HA1)

#### Lack of quality assurance

Controlling and improving anaesthesia quality is crucial, but impossible without sufficient qualified anaesthetists. All participants indicated that quality cannot be imposed unless it is habituated. However, there appeared little incentive. Most patients never considered questioning anaesthetists’ qualifications the way they did surgeons nor did anaesthetists consider complaining formally about insufficient or outdated equipment.

> *“As Aristotle said, ‘Quality is a not an act it’s a habit!’ So one institution or person cannot impose it unless everyone makes it a habit. HCC can only act on a complaint. Why must only patients complain in case of mishaps? Why don’t doctors ever complain in cases of outdated and insufficient equipment? Why don’t anaesthetists complain if they notice unqualified personal, or worn-out equipment in a hospital? Why do they risk patients’ lives?” (HC1)*

Most participants noted that once HCC licenced a hospital, it only visited to check on complaints or for periodic availability monitoring of essential facilities and equipment. HCC required that hospital quality improvement committees conduct all quality assurance. Many considered this ‘self-monitoring’ approach unreliable.

> “*the healthcare commission in Pakistan has minimum standards for practice, but they do not check quality, i*.*e*., *their standard is just to have anaesthesia machine, pulse oximeter, but no emphasis on quality of this machinery […]. They haven’t specified the type and quality and maintenance…” (HA9)*

#### Pre-operative and intra-operative training gaps

Some quality-of-care challenges related to insufficient training. All described continuous medical education (CME) as both insufficient and underused, with peripheral anaesthetists working for decades without updating their knowledge or skills.

> “*In the entire world, medical councils don’t renew licences for doctors until they have certification of recent CME credit hours. PMDC [Pakistan medical and dental council] started this in 2016 and faced immense backlash. Unfortunately, we want perks of working abroad but not legislation like them*.*” (HC3)*

Several complained that both CME trainers and trainees expected financial incentives for participating.

> “*It’s for their own sake. How can a graduate from 1980 work on 2021 standards without updating his knowledge? They need to learn newer techniques to improve their practice and it’s an incentive that our government is providing it free of charge, but they all want monetary incentives even to take CME!”* (HD5)

However, others clarified that CME imposed an additional burden on already overwhelmed anaesthesiologists and was therefore resisted.

> *“One of the main reasons for minimal interest is it’s imposed alongside regular work, so it overburdens the already burdened physicians. CME should be given in 2 phases, half attend training while other half works, or people from tertiary care substitute for them so they can attend training with a free mind*…” (HA9)

As a senior participant added:

> “*Government don’t even give incentives to CME providers. If I’m invited to give training I would go at this age, but a young consultant managing a private practice would not be interested in providing CME training without incentive as he would be compromising his private practice and patients*.” (HA10)

Many participants insisted on the urgency of training doctors at frontline facilities in handling trauma and emergency fluid resuscitation, as requiring airway clearing and starting resuscitation for trauma cases at primary level and ensuring availability of pulse oximeters could save many lives.

*“The main reason for increased disease burden and mortality is lack of emergency handling at basic level. A doctor in BHU [basic health unit] or RHC [rural health centre] could start resuscitation or clear airways in trauma cases before sending to bigger hospitals. This could be lifesaving, as usually when such trauma patients reach THQ or DHQ, they have lost so much fluid that saving them is impossible. It’s basic training that should be required before entering the system*.*” (HC1)*

### Discrepancies between specialities

We identified two sub-themes affecting inter-specialties discrepancies: (i) surgeon dominance; (ii) mixed perceptions on allied anaesthesia staff.

#### Surgeon dominance

Most participants mentioned inter-speciality disparities as challenging anaesthesia delivery. For example, several suggested the reason Punjab had 800 posts for consultant surgeons and only 50-70 for consultant anaesthesiologists was that surgeons held the top posts and promoted colleagues from their speciality.

> *“Anaesthetists are masked by their surgery and medicine colleagues who never let their problems be heard. I have never seen any anaesthetist as [senior management]. These usually belong to medicine or surgery and that’s what we hear about all the time. Their problems, their seats, their betterment*… *Doctors need to work together for the betterment of the health department!” (HD2)*

Others noted similar discrepancies in teaching hospitals and universities.

*“…medical personnel sitting in higher posts just promotes his own specialty. Same with professorship. In gynaecology surgery we have 30-40 professors, but anaesthesia just has 9-10*.” (HD7)

Participants suggested this lack of policy-making power and influence contributed to increased anaesthetist workload and reliance on unqualified personnel, both of which affected quality.

#### Mixed perceptions on allied anaesthesia staff

Participants described intra-specialty discrepancies, sharing mixed views on the role of allied anaesthesia professionals (e.g. anaesthesia assistants, nurse-anaesthetists, anaesthesia technologists with 2-4 years of training respectively). Health department participants were strongly in favour of allied anaesthesia staff, as they could reduce workload and nurse-anaesthetists had demonstrated their effectiveness in army hospitals nationwide.

> *“Nurse-anaesthetists are working very effectively in army hospitals due to strict checks and balances. If it can be maintained in one sector, why not in others? The only reason is insecurity of seniors*…*”* (HD2)
>
> *“Government has allowed doctors with training in anaesthesia (FCPS/MCPS/MS) who somehow have not passed the exam, to practice aesthesia freely to overcome the severe shortage in the system. The allied anaesthesia workforce will increase effectiveness and minimize risks…” (HC5)*

Despite claims of anaesthesiologists’ ‘insecurity’ towards allied anaesthesia professionals, the primary concern appeared to be that they often functioned without sufficient supervision and monitoring.

> *“Sadly, we’re producing practically helpful people, but improper practice guidelines are turning them into havoc*.” (HA3)

Several professors condemned the reliance on allied professionals, clarifying concerns about lack of guidance and monitoring.

“*Technologists or nurse-anaesthetists are not well trained in handling emergencies. They must work under the supervision of consultants. However, the lack of checks and balances and demand for low-wage workers promotes their solo private practice, which decreases health system quality instead of improving it*.*” (HA8)*

Some noted that nurse-anaesthetists and anaesthesia technologists were more useful than MOs, who were not specialists and rotated departments while allied professionals could not be moved and had 2-4 years specialised training.

> “*Technologists could be even better placed than MOs, as in DHQ/THQ they change MOs every few months. At least 3 months are required to train an MO enough in anaesthesia that he can take care of a patient alone in theatre. When finally he is trained, he is rotated to some other department and a new one is provided with zero experience, which is an utter waste of time and energy*. (HA6)

Many suggested that an effective legal framework, technical guidance, and oversight would ensure the beneficial participation of allied professionals.

“*To reduce the workload in peripheral areas, every consultant must be attached with 3-4 technologists or nurse-anaesthetists. This would not only decrease the workload in peripheral areas but also limit their unsupervised practice*.*”* (HA9)

## DISCUSSION

This study is the first to highlight on-going challenges within the anaesthesia delivery system in Punjab province as experienced by senior practitioners. Challenges described in recruitment and retention, quality-of-care, training, and inter/intra-specialty discrepancies align with findings in several African LMICS, including Malawi, Rwanda, and Uganda (22-24).

### Anaesthetist recruitment and retention

Loss of skilled staff, due to emigration, burnout, or gender-related barriers, was a central challenge to providing quality anaesthesia services in Punjab. Both academic and government participants described significant numbers being trained and qualifying every year but not joining the system, as noted in other LMICs (25). This increased the workload for those already in the system, compounded by increasing requirements from rapid surgical improvements, resulting in considerable burnout (26) or dissatisfaction (27). Maldistribution was similarly noted in other settings (28). Recruitment challenges and ‘brain drain’ to HICs also aligned with the literature (29). The significant numbers of women consultants not actively participating in the workforce is a recognised phenomenon in Pakistan and noted for other specialties (30, 31).

Addressing these systemic issues requires political will. A first step may be requiring multi-year bonds for government service after graduation, mandatory peripheral rotations to improve equity, and addressing barriers for women anaesthetists. For example, consultant-anaesthetists could be required to work in-country for 3-5 years while allied professionals could be posted peripherally under supervision of designated consultants to increase anaesthesia services capacity and quality. Further research is needed to identify the most effective ways to improve the recruitment and retention of women clinicians and allied professionals in the health system.

### Quality-of-care

Pakistan’s HCC provides minimum standards for anaesthesia practice in accordance with international guidelines (32), but as noted in our findings this quantifies availability of essential equipment and medication while quality and maintenance remains a hospital responsibility. This works for hospitals with robust internal systems, but the lack of follow-up and external accountability is worrisome (4, 33). Quality is a two-way process, and practicing anaesthetists must feel empowered to report it to HCC for necessary actions if quality is not maintained by hospitals, as this will affect their patients (34). However, our findings suggested a reason for the absence of focus on anaesthesia quality is lack of public awareness - as found in other Asian literature - with patients looking for qualified surgeons but not considering qualified anaesthetists, enabling under-qualified anaesthetists to continue practicing (35-37). Moreover, practice outside theatre would allow greater patient interaction during pain management but this remains limited for anaesthetists in Punjab due to competition from other specialities and lack of training (2, 38).

Addressing these issues requires a combination of mandatory but feasible essential CME to upgrade decades-old skills among peripheral consultants and public engagement about the broad benefits and importance of anaesthesiology, along with increased retention so more anaesthesiologists have time to seek hospital leadership roles or raise awareness of the specialty.

### Discrepancies between specialities

As surgery and anaesthesia work together, numbers of surgical and anaesthesiology residents, doctors, and consultants should be proportional to manage workloads and avoid burnout (27). However, anaesthesia is less visible - described by several participants as “*behind the curtain*” and faces major staffing shortages even in urban areas. Inter-specialty discrepancies are evident in Pakistan, as in many LMICs, which affect anaesthesia staffing and equipment investments and thus reduce quality and safety (4,5,26). This is worsened in peripheral areas as non-physician anaesthetists are not allowed to practice due to insufficient legislation and ongoing biases (39,40). Although allied health professionals have participated effectively in anaesthesia provision elsewhere, their numbers in Punjab are negligible (4).

Working to address these discrepancies will require developing clear national legislation and practice guidelines for allied health professionals and, as with quality-of-care, raising the public profile of anaesthesia in Pakistan.

### Limitations

Several limitations should be considered. First, only senior perspectives were included, and frontline or patient perspectives might have provided additional insights. Second, participants were all from Punjab province, which has relatively high socioeconomic, educational, and health provision indicators and anaesthesia provision in other provinces is potentially worse. Third, the sample is relatively small, though we did aim for data saturation, and some nuances may have been missed.

## CONCLUSIONS

Improving anaesthesia delivery in Punjab will require increasing recruitment, retention, and peripheral distribution of physician and non-physician anaesthetists. This may require 3-5 year bonds for government service post-graduation, addressing barriers for women anaesthetists, and mandatory peripheral rotations to improve equity. Continuous medical education must be mandatory but also practical (e.g. at appropriate times and locations, with sufficient incentives so anaesthetists do not lose income by attending).

## Data Availability

An anonymised and de-identified dataset can be accessed through the University of the Punjab repository via the lead author.

## DECLARATIONS

### Conflicts of interest

None declared.

## Acknowledgements

We would like to thank all study participants for sharing their time and experience.

## Author contributions

SS conceived the study with support from RZ. SS collected and analysed data and drafted the manuscript with supervision by NH. NH contributed to analysis and interpretation and critically revised the manuscript. All authors reviewed and approved the version for submission.

## Funding

None received.

## Notes

### Competing Interest Statement

The authors have declared no competing interest.

### Clinical Trial

NA

### Funding Statement

No funding was provided.

### Author Declarations

The University of the Punjab Department of Public Health Doctoral Program Committee provided study approval (D/154/ISCS March 18, 2021).

